# Hemorrhagic Transformation After Endovascular Thrombectomy in Young Adults: A Prediction Model

**DOI:** 10.64898/2026.06.03.26354874

**Authors:** Qiushi Lv, Kang Yuan, Anyu Liao, Zhaojun Wang, Yun Li, Guodong Xiao, Wenhua Liu, Zhiming Zhou, Dong Yang, Kangmo Huang, Can Chen, Weichen Dong, Liangyuan Pan, Wusheng Zhu, Xinfeng Liu

## Abstract

**Background and Purpose:** Hemorrhagic transformation (HT) is a serious complication of endovascular thrombectomy (EVT), yet dedicated prediction models for young adults are lacking. We aimed to develop and externally validate a simplified risk score for HT in young adults with acute ischemic stroke undergoing EVT.

**Methods:** This multicenter retrospective study included patients aged 18 to 49 years with acute anterior circulation large vessel occlusion who underwent EVT. The primary outcome was any HT within 24 hours after EVT. Multivariable logistic regression was used to identify independent predictors of HT, from which the NO-PAIN Score was derived. External validation was performed in an independent cohort of 138 patients.

**Results:** Among 598 patients in the derivation cohort, HT occurred in 176 (29.4%). Five independent predictors were identified: admission NIHSS, number of thrombectomy passes, atrial fibrillation, alcohol consumption, and mTICI grade. The mTICI grade demonstrated a non-linear, inverted U-shaped relationship with HT risk, peaking at partial recanalization. The NO-PAIN Score showed acceptable discrimination in both the derivation (C-index, 0.737; optimism-corrected C-index, 0.748) and external validation cohorts (C-index, 0.726), with satisfactory calibration.

**Conclusions:** The NO-PAIN Score is a simple risk prediction tool for HT after EVT in young adults with acute anterior circulation large vessel occlusion. It may assist in individualized risk stratification in this population.

## Introduction

Ischemic stroke in young adults, defined as occurring in individuals aged 18 to 49 years, accounts for 10% to 20% of all ischemic strokes, and its incidence continues to rise globally.^1,2^ Compared with older patients, young stroke survivors face a longer lifetime burden of disability and a greater socioeconomic impact, as many are primary breadwinners for their families.^3^ Endovascular thrombectomy (EVT) has become the standard of care for acute large vessel occlusion. Recent registry data indicate that compared with older patients, young patients achieve better functional outcomes after EVT,^4,5^ yet approximately one third still fare poorly at 90 days.^5^ These studies have focused predominantly on functional outcomes, while the risk profile for periprocedural complications remains limited.

Hemorrhagic transformation (HT) is one of the most serious complications of EVT, occurring in 25% to 45% of treated patients.^6-8^ Both symptomatic intracranial hemorrhage (sICH) and asymptomatic HT have been associated with worse functional outcomes and higher mortality after EVT.^9^⁻^11^ Several prediction models have been developed specifically for sICH after EVT, including the ASIAN score, the IER-SICH nomogram, and the TREAT-AIS score.^12^⁻^14^ However, these models were developed predominantly in older or mixed-age cohorts, and were designed to predict sICH rather than any radiographic HT. A dedicated prediction model for HT in young adults undergoing EVT has not been established.

This study aimed to develop and externally validate a simplified risk score, the NO-PAIN Score, for predicting HT in young adults after EVT for acute anterior circulation large vessel occlusion.

## Methods

### Study Design

This multicenter retrospective study was approved by the institutional review boards of all participating centers, with the coordinating center (Affiliated Jinling Hospital, School of Medicine, Nanjing University) providing the primary ethical approval (reference number: 2023DZKY-125-01). The requirement for written informed consent was waived owing to the retrospective design. The study adhered to the STROBE guidelines.^15^

### Data Source and Setting

Patients were identified from prospectively maintained stroke registries at four comprehensive stroke centers in China between January 2017 and December 2023 (Table S1). The external validation cohort was derived from three independent sources: the ACTUAL study,^16^ the Captor trial (ChiCTR1900025256, https://www.chictr.org.cn), and the SINOMED SR trial (NCT04973332, clinicaltrials.gov). From these sources, we extracted all young adult patients with acute anterior circulation large vessel occlusion who underwent EVT.

### Study Population

Inclusion criteria were age 18 to 49 years, acute anterior circulation large vessel occlusion, and EVT within 24 hours of symptom onset. Intravenous thrombolysis before EVT was permitted. We excluded patients with posterior circulation occlusion, pre-stroke modified Rankin Scale (mRS) score >2, missing critical data or loss to follow-up, or pre-EVT intracranial hemorrhage.

### Outcomes

The primary outcome was any HT, assessed on follow-up neuroimaging within 24 hours after EVT and classified according to the Heidelberg Bleeding Classification.^17^ This classification distinguishes hemorrhagic infarction (HI1, HI2) from parenchymal hematoma (PH1, PH2), and further defines remote parenchymal hematoma, intraventricular hemorrhage, subarachnoid hemorrhage, and subdural hemorrhage as distinct categories. The secondary outcome was sICH, defined as radiographic hemorrhage plus neurological deterioration (NIHSS increase >4 points, or >2 points in any single category, or requiring major intervention). All neuroimaging studies were independently reviewed by two neuroradiologists blinded to clinical outcomes, with disagreements resolved by consensus or by a third reader.

### Covariates

We collected data on demographic characteristics, vascular risk factors, clinical variables, procedural details, and laboratory parameters. The neutrophil-to-lymphocyte ratio (NLR), platelet-to-lymphocyte ratio (PLR), and systemic inflammatory response index (SIRI) were calculated as: NLR = neutrophil/lymphocyte count; PLR = platelet/lymphocyte count; and SIRI = (neutrophil × monocyte) / lymphocyte count. Collateral circulation was assessed using the American Society of Interventional and Therapeutic Neuroradiology/Society of Interventional Radiology (ASITN/SIR) grading system, with grade 0 to 1 indicating poor, grade 2 indicating moderate, and grade 3 to 4 indicating good collateral status.^18^ The degree of recanalization was assessed using the modified Thrombolysis in Cerebral Infarction (mTICI) scale, with successful recanalization defined as mTICI 2b or 3.

### Statistical Analysis

The derivation cohort included 598 patients, of whom 176 (29.4%) developed HT. Based on the commonly recommended rule of 10 events per candidate predictor variable, this sample permitted the inclusion of approximately 17 variables in the final multivariable model. Missing data were infrequent across all baseline variables (<5% for each). Given the low proportion of missingness, complete-case analysis was performed without imputation.

To reduce detection bias, all outcome assessments (HT and sICH) were performed by neuroradiologists blinded to clinical outcomes. Selection bias was minimized by including consecutive eligible patients from prospective registries at four centers. Information bias was mitigated by using standardized definitions for all variables and automated laboratory measurements. Potential confounding was adjusted in multivariable logistic regression models, and internal validation with 1000 bootstrap resamples was performed to assess model stability.

Continuous variables are presented as mean ± standard deviation or median (interquartile range) and were compared using the t-test or Mann-Whitney U test. Categorical variables are presented as frequency (percentage) and were compared using the χ² test or Fisher exact test. Variables with P<0.10 in univariable analysis were entered into multivariable logistic regression with backward stepwise selection. Both the mTICI grade and collateral circulation grade were modeled as ordinal variables with polynomial contrasts to capture potential nonlinear associations. Model discrimination was assessed using the area under the receiver operating characteristic curve (AUC). Calibration was evaluated with the Hosmer-Lemeshow test and a calibration plot. A simplified point-score system was constructed from the final model. For external validation, the same model formula and coefficients were applied to the external cohort. A two-tailed P<0.05 was considered significant. All analyses were performed using R version 4.5.2.

## Results

### Study Population and Baseline Characteristics

The derivation cohort included 598 patients with acute anterior circulation large vessel occlusion who underwent EVT (Figure S1A). Their median age was 46 years (IQR, 42-48), 77.9% were male, and the median admission NIHSS was 13 (IQR, 10-18). HT occurred in 176 patients (29.4%) and sICH in 68 (11.4%). Baseline characteristics of the cohort, grouped by HT and sICH status, are presented in Tables 1 and 2.

**Table 1.**
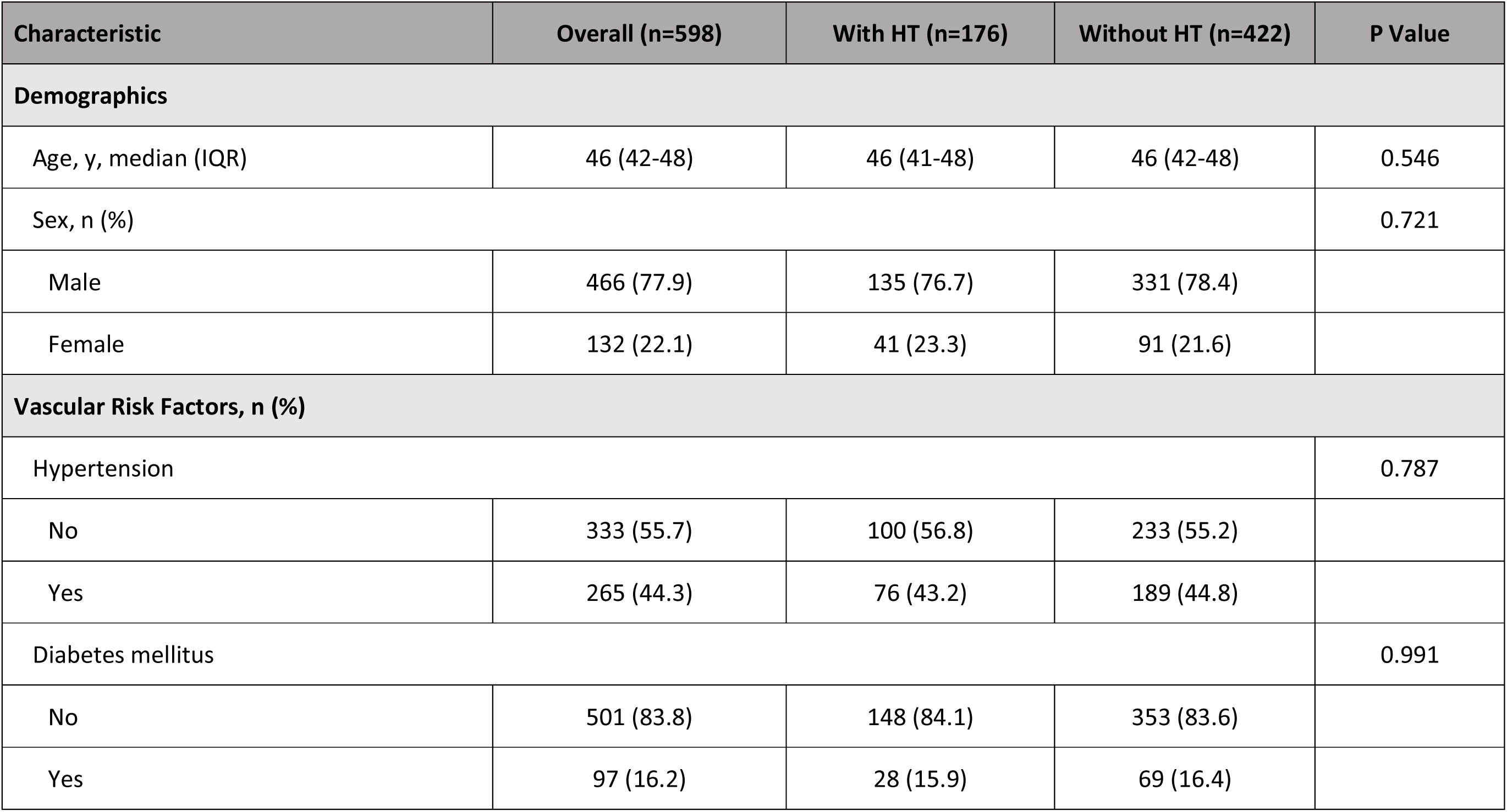
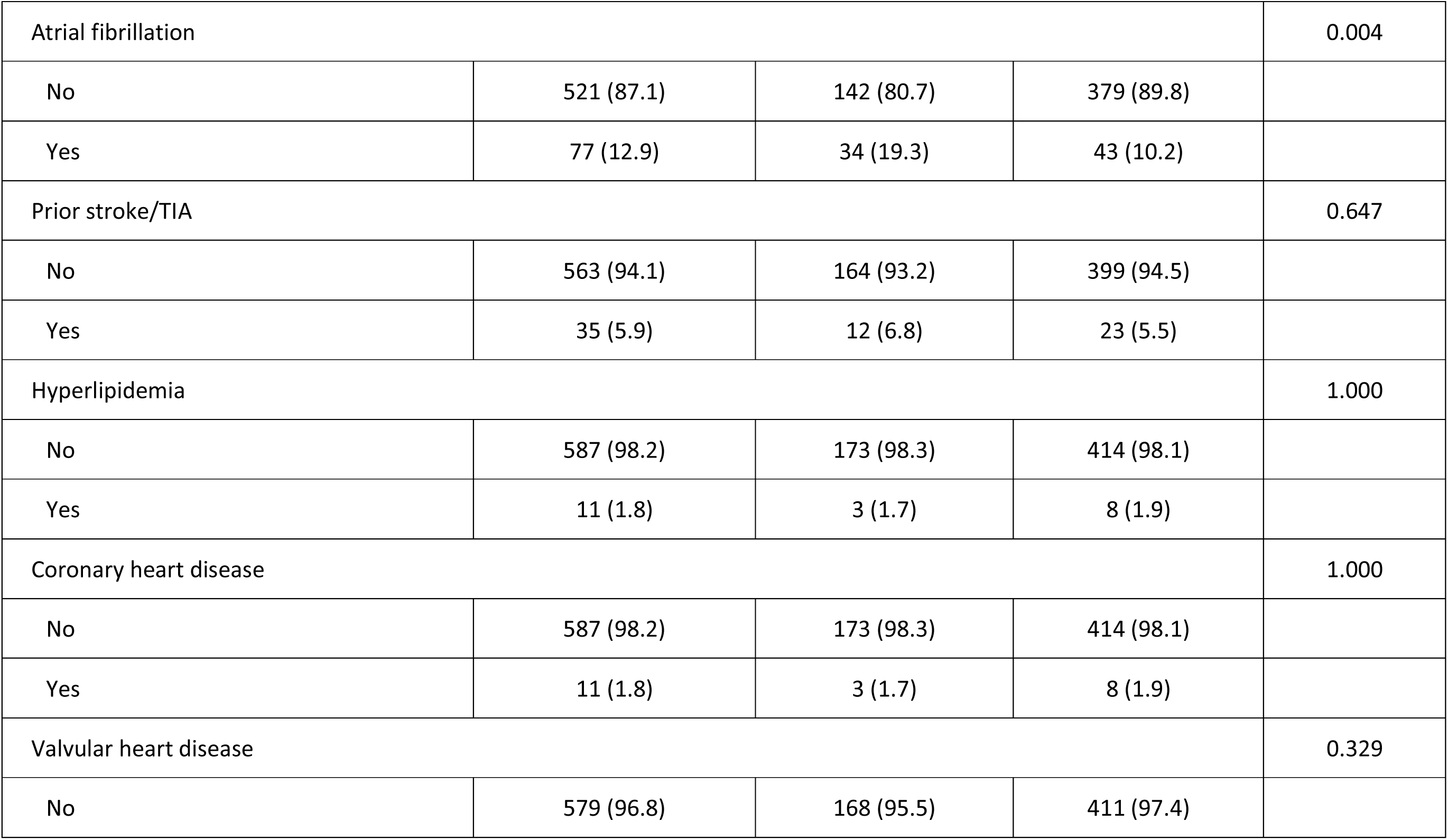
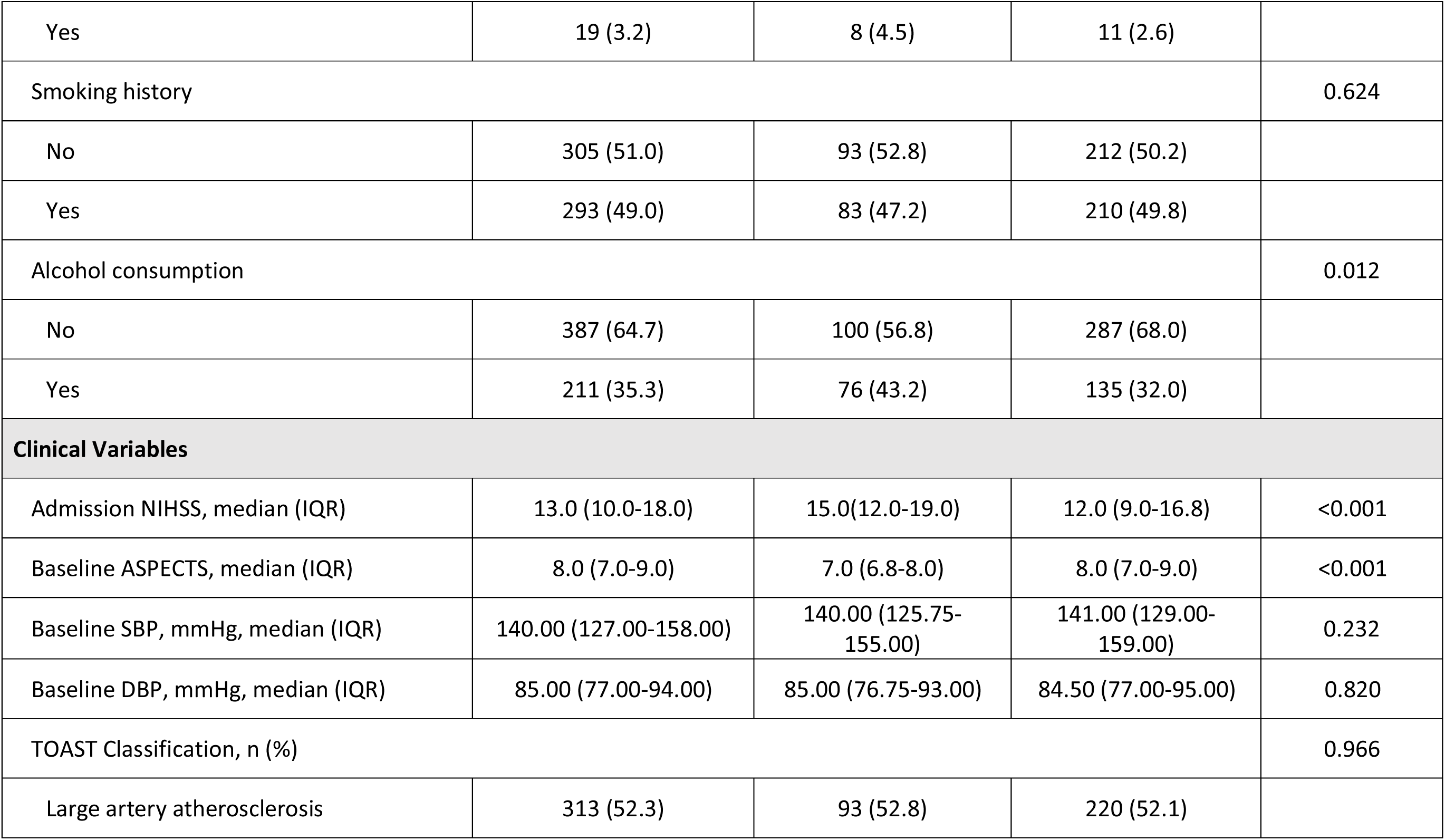
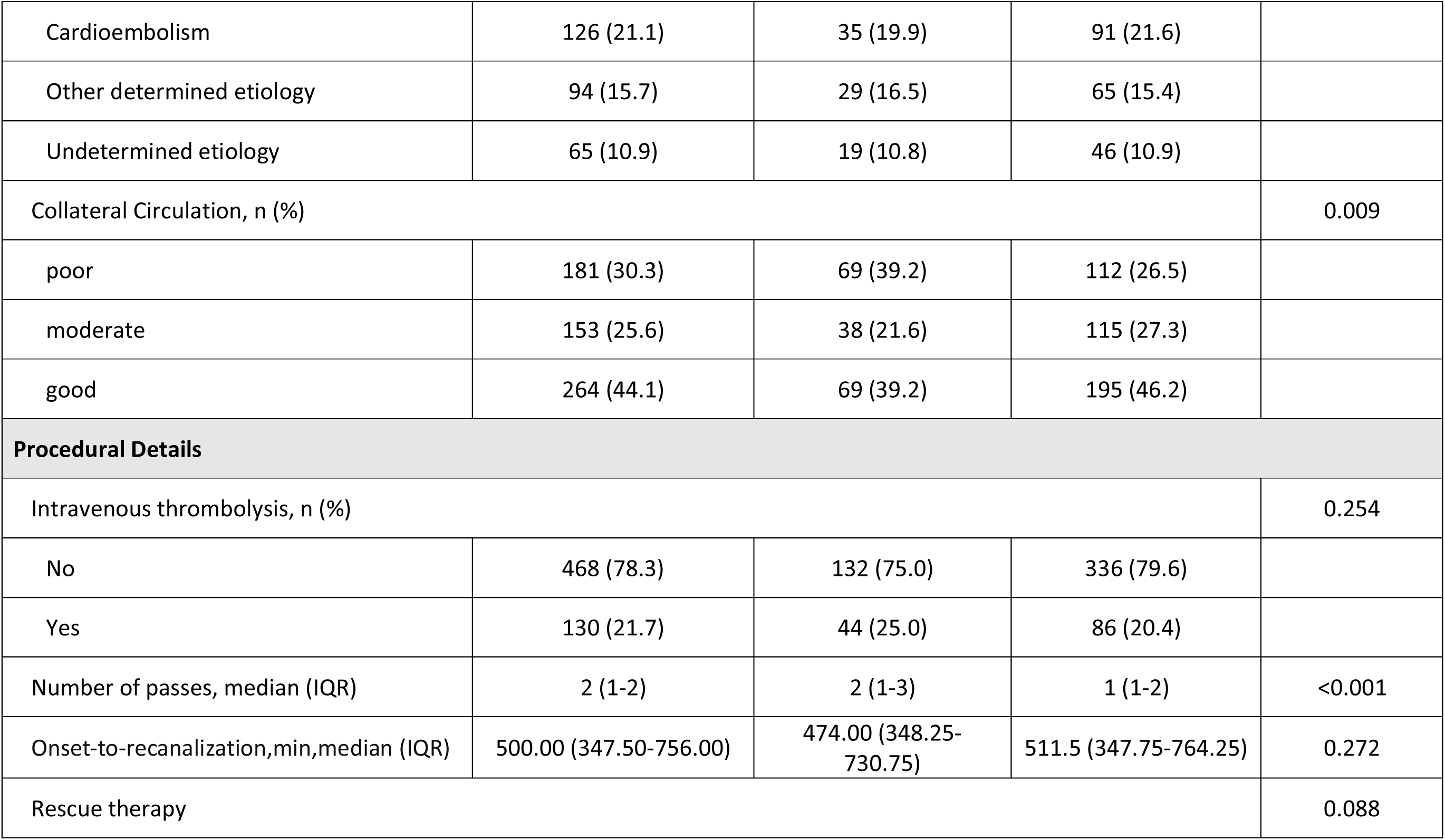
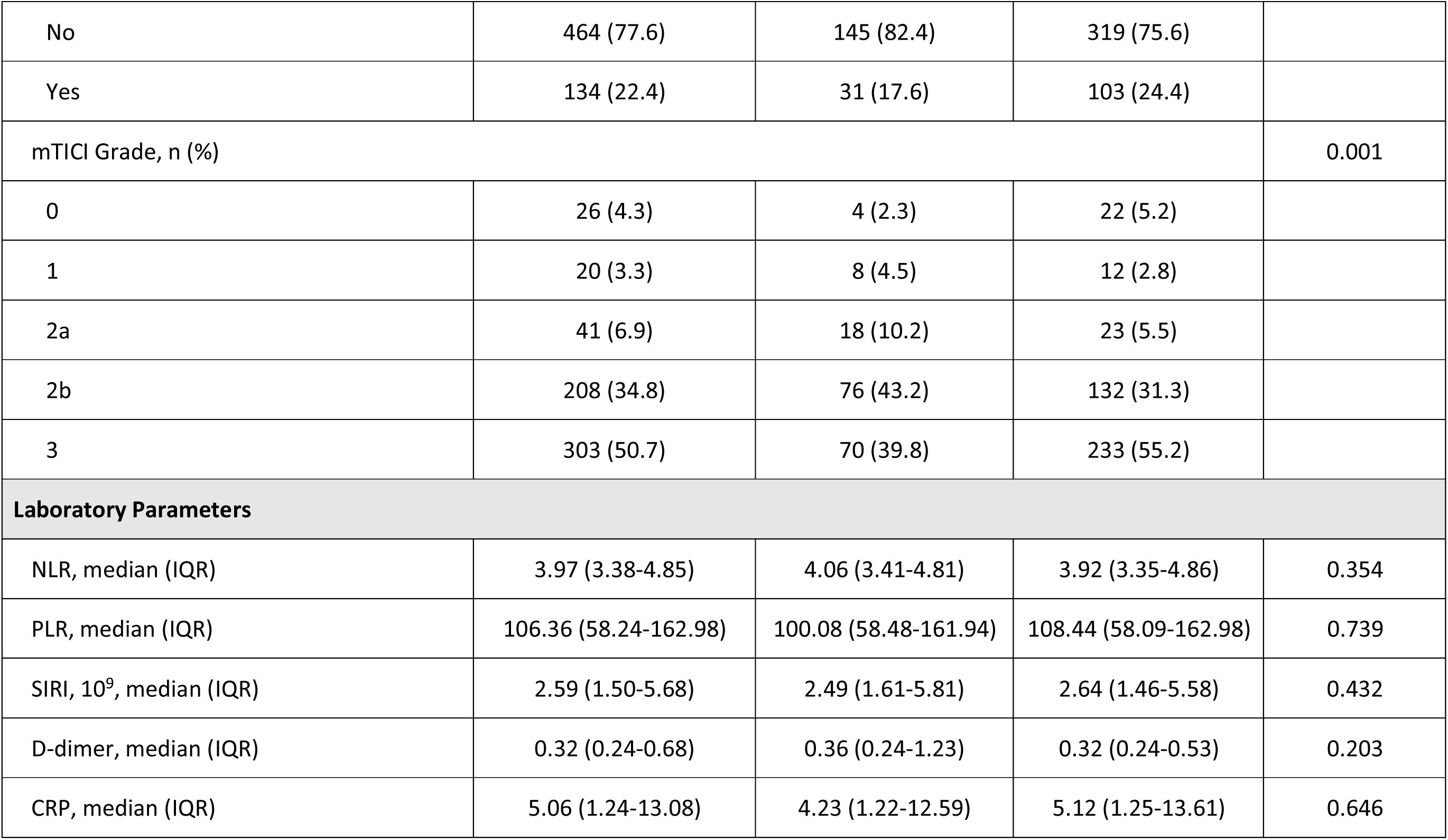
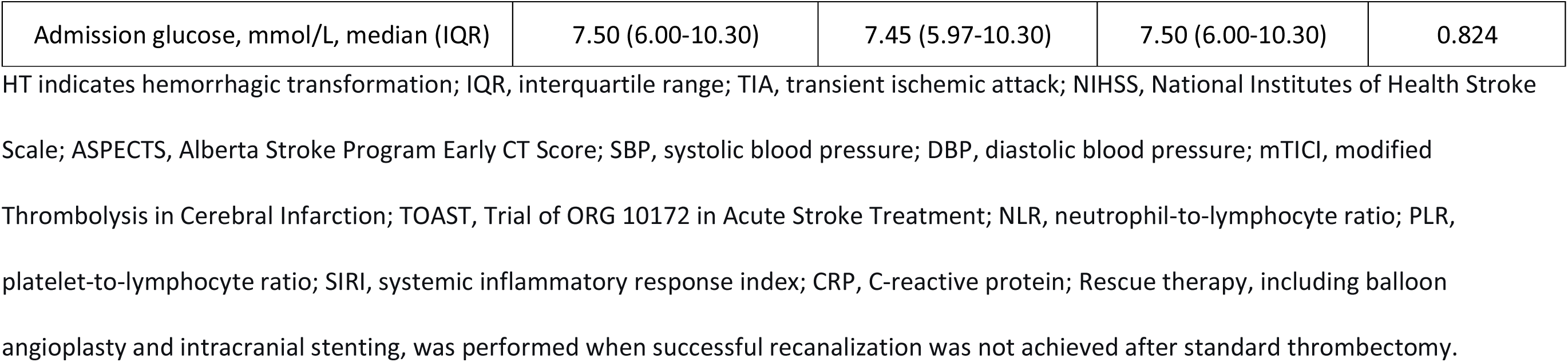
Baseline Characteristics of the Derivation Cohort, Stratified by Hemorrhagic Transformation.

| Characteristic | Overall (n=598) | With HT (n=176) | Without HT (n=422) | P Value |
| --- | --- | --- | --- | --- |
| **Demographics** |  |  |  |  |
| Age, y, median (IQR) | 46 (42-48) | 46 (41-48) | 46 (42-48) | 0.546 |
| Sex, n (%) |  |  |  | 0.721 |
| Male | 466 (77.9) | 135 (76.7) | 331 (78.4) |  |
| Female | 132 (22.1) | 41 (23.3) | 91 (21.6) |  |
| **Vascular Risk Factors, n (%)** |  |  |  |  |
| Hypertension |  |  |  | 0.787 |
| No | 333 (55.7) | 100 (56.8) | 233 (55.2) |  |
| Yes | 265 (44.3) | 76 (43.2) | 189 (44.8) |  |
| Diabetes mellitus |  |  |  | 0.991 |
| No | 501 (83.8) | 148 (84.1) | 353 (83.6) |  |
| Yes | 97 (16.2) | 28 (15.9) | 69 (16.4) |  |
| Atrial fibrillation |  |  |  | 0.004 |
| No | 521 (87.1) | 142 (80.7) | 379 (89.8) |  |
| Yes | 77 (12.9) | 34 (19.3) | 43 (10.2) |  |
| Prior stroke/TIA |  |  |  | 0.647 |
| No | 563 (94.1) | 164 (93.2) | 399 (94.5) |  |
| Yes | 35 (5.9) | 12 (6.8) | 23 (5.5) |  |
| Hyperlipidemia |  |  |  | 1.000 |
| No | 587 (98.2) | 173 (98.3) | 414 (98.1) |  |
| Yes | 11 (1.8) | 3 (1.7) | 8 (1.9) |  |
| Coronary heart disease |  |  |  | 1.000 |
| No | 587 (98.2) | 173 (98.3) | 414 (98.1) |  |
| Yes | 11 (1.8) | 3 (1.7) | 8 (1.9) |  |
| Valvular heart disease |  |  |  | 0.329 |
| No | 579 (96.8) | 168 (95.5) | 411 (97.4) |  |
| Yes | 19 (3.2) | 8 (4.5) | 11 (2.6) |  |
| Smoking history |  |  |  | 0.624 |
| No | 305 (51.0) | 93 (52.8) | 212 (50.2) |  |
| Yes | 293 (49.0) | 83 (47.2) | 210 (49.8) |  |
| Alcohol consumption |  |  |  | 0.012 |
| No | 387 (64.7) | 100 (56.8) | 287 (68.0) |  |
| Yes | 211 (35.3) | 76 (43.2) | 135 (32.0) |  |
| **Clinical Variables** |  |  |  |  |
| Admission NIHSS, median (IQR) | 13.0 (10.0-18.0) | 15.0(12.0-19.0) | 12.0 (9.0-16.8) | <0.001 |
| Baseline ASPECTS, median (IQR) | 8.0 (7.0-9.0) | 7.0 (6.8-8.0) | 8.0 (7.0-9.0) | <0.001 |
| Baseline SBP, mmHg, median (IQR) | 140.00 (127.00-158.00) | 140.00 (125.75-155.00) | 141.00 (129.00-159.00) | 0.232 |
| Baseline DBP, mmHg, median (IQR) | 85.00 (77.00-94.00) | 85.00 (76.75-93.00) | 84.50 (77.00-95.00) | 0.820 |
| TOAST Classification, n (%) |  |  |  | 0.966 |
| Large artery atherosclerosis | 313 (52.3) | 93 (52.8) | 220 (52.1) |  |
| Cardioembolism | 126 (21.1) | 35 (19.9) | 91 (21.6) |  |
| Other determined etiology | 94 (15.7) | 29 (16.5) | 65 (15.4) |  |
| Undetermined etiology | 65 (10.9) | 19 (10.8) | 46 (10.9) |  |
| Collateral Circulation, n (%) |  |  |  | 0.009 |
| poor | 181 (30.3) | 69 (39.2) | 112 (26.5) |  |
| moderate | 153 (25.6) | 38 (21.6) | 115 (27.3) |  |
| good | 264 (44.1) | 69 (39.2) | 195 (46.2) |  |
| **Procedural Details** |  |  |  |  |
| Intravenous thrombolysis, n (%) |  |  |  | 0.254 |
| No | 468 (78.3) | 132 (75.0) | 336 (79.6) |  |
| Yes | 130 (21.7) | 44 (25.0) | 86 (20.4) |  |
| Number of passes, median (IQR) | 2 (1-2) | 2 (1-3) | 1 (1-2) | <0.001 |
| Onset-to-recanalization,min,median (IQR) | 500.00 (347.50-756.00) | 474.00 (348.25-730.75) | 511.5 (347.75-764.25) | 0.272 |
| Rescue therapy |  |  |  | 0.088 |
| No | 464 (77.6) | 145 (82.4) | 319 (75.6) |  |
| Yes | 134 (22.4) | 31 (17.6) | 103 (24.4) |  |
| mTICI Grade, n (%) |  |  |  | 0.001 |
| 0 | 26 (4.3) | 4 (2.3) | 22 (5.2) |  |
| 1 | 20 (3.3) | 8 (4.5) | 12 (2.8) |  |
| 2a | 41 (6.9) | 18 (10.2) | 23 (5.5) |  |
| 2b | 208 (34.8) | 76 (43.2) | 132 (31.3) |  |
| 3 | 303 (50.7) | 70 (39.8) | 233 (55.2) |  |
| **Laboratory Parameters** |  |  |  |  |
| NLR, median (IQR) | 3.97 (3.38-4.85) | 4.06 (3.41-4.81) | 3.92 (3.35-4.86) | 0.354 |
| PLR, median (IQR) | 106.36 (58.24-162.98) | 100.08 (58.48-161.94) | 108.44 (58.09-162.98) | 0.739 |
| SIRI, 10<sup>9</sup>, median (IQR) | 2.59 (1.50-5.68) | 2.49 (1.61-5.81) | 2.64 (1.46-5.58) | 0.432 |
| D-dimer, median (IQR) | 0.32 (0.24-0.68) | 0.36 (0.24-1.23) | 0.32 (0.24-0.53) | 0.203 |
| CRP, median (IQR) | 5.06 (1.24-13.08) | 4.23 (1.22-12.59) | 5.12 (1.25-13.61) | 0.646 |
| Admission glucose, mmol/L, median (IQR) | 7.50 (6.00-10.30) | 7.45 (5.97-10.30) | 7.50 (6.00-10.30) | 0.824 |

HT indicates hemorrhagic transformation; IQR, interquartile range; TIA, transient ischemic attack; NIHSS, National Institutes of Health Stroke Scale; ASPECTS, Alberta Stroke Program Early CT Score; SBP, systolic blood pressure; DBP, diastolic blood pressure; mTICI, modified Thrombolysis in Cerebral Infarction; TOAST, Trial of ORG 10172 in Acute Stroke Treatment; NLR, neutrophil-to-lymphocyte ratio; PLR, platelet-to-lymphocyte ratio; SIRI, systemic inflammatory response index; CRP, C-reactive protein; Rescue therapy, including balloon angioplasty and intracranial stenting, was performed when successful recanalization was not achieved after standard thrombectomy.

**Table 2.** Baseline Characteristics of the Derivation Cohort, Stratified by Symptomatic Intracranial Hemorrhage.

| Characteristic | Overall (n=598) | with sICH (n=68) | without sICH (n=530) | P Value |
| --- | --- | --- | --- | --- |
| **Demographics** |  |  |  |  |
| Age, y, median (IQR) | 46 (42-48) | 46 (43-48) | 46 (42-48) | 0.689 |
| Sex, n (%) |  |  |  | 0.161 |
| Male | 466 (77.9) | 58 (85.3) | 408 (77.0) |  |
| Female | 132 (22.1) | 10 (14.7) | 122 (23.0) |  |
| **Vascular Risk Factors, n (%)** |  |  |  |  |
| Hypertension |  |  |  | 0.258 |
| No | 333 (55.7) | 33 (48.5) | 300 (56.6) |  |
| Yes | 265 (44.3) | 35 (51.5) | 230 (43.4) |  |
| Diabetes mellitus |  |  |  | 0.608 |
| No | 501 (83.8) | 55 (80.9) | 446 (84.2) |  |
| Yes | 97 (16.2) | 13 (19.1) | 84 (15.8) |  |
| Atrial fibrillation |  |  |  | 0.291 |
| No | 521 (87.1) | 56 (82.4) | 465 (87.7) |  |
| Yes | 77 (12.9) | 12 (17.6) | 65 (12.3) |  |
| Prior stroke/TIA |  |  |  | 0.167 |
| No | 563 (94.1) | 61 (89.7) | 502 (94.7) |  |
| Yes | 35 (5.9) | 7 (10.3) | 28 (5.3) |  |
| Hyperlipidemia |  |  |  | 0.472 |
| No | 587 (98.2) | 68 (100.0) | 519 (97.9) |  |
| Yes | 11 (1.8) | 0 (0.0) | 11 (2.1) |  |
| Coronary heart disease |  |  |  | 0.472 |
| No | 587 (98.2) | 68 (100.0) | 519 (97.9) |  |
| Yes | 11 (1.8) | 0 (0.0) | 11 (2.1) |  |
| Valvular heart disease |  |  |  | 0.223 |
| No | 579 (96.8) | 68 (100.0) | 511 (96.4) |  |
| Yes | 19 (3.2) | 0 (0.0) | 19 (3.6) |  |
| Smoking history |  |  |  | 0.214 |
| No | 305 (51.0) | 40 (58.8) | 265 (50.0) |  |
| Yes | 293 (49.0) | 28 (41.2) | 265 (50.0) |  |
| Alcohol consumption |  |  |  | 0.499 |
| No | 387 (64.7) | 41 (60.3) | 346 (65.3) |  |
| Yes | 211 (35.3) | 27 (39.7) | 184 (34.7) |  |
| **Clinical Variables** |  |  |  |  |
| Admission NIHSS, median (IQR) | 13.0 (10.0-18.0) | 16.0 (13.0-20.0) | 12.0 (10.0-17.0) | <0.001 |
| Baseline ASPECTS, median (IQR) | 8.0 (7.0-9.0) | 7.0 (6.0-9.0) | 8.0 (7.0-9.0) | 0.006 |
| Baseline SBP, mmHg, median (IQR) | 140.00 (127.00-158.00) | 140.00 (125.75-160.00) | 140.00 (128.00-156.00) | 0.730 |
| Baseline DBP, mmHg, median (IQR) | 85.00 (77.00-94.00) | 86.00 (76.00-93.50) | 85.00 (77.00-94.00) | 0.695 |
| **Collateral Circulation, n (%)** |  |  |  | <0.001 |
| poor | 181 (30.3) | 37 (54.4) | 144 (27.2) |  |
| moderate | 153 (25.6) | 10 (14.7) | 143 (27.0) |  |
| good | 264 (44.1) | 21 (30.9) | 243 (45.8) |  |
| **TOAST Classification, n (%)** |  |  |  | 0.790 |
| Large artery atherosclerosis | 313 (52.3) | 36 (52.9) | 277 (52.3) |  |
| Cardioembolism | 126 (21.1) | 16 (23.5) | 110 (20.8) |  |
| Other determined etiology | 94 (15.7) | 8 (11.8) | 86 (16.2) |  |
| Undetermined etiology | 65 (10.9) | 8 (11.8) | 57 (10.8) |  |
| **Procedural Details** |  |  |  |  |
| Intravenous thrombolysis, n (%) |  |  |  | 0.930 |
| No | 468 (78.3) | 54 (79.4) | 414 (78.1) |  |
| Yes | 130 (21.7) | 14 (20.6) | 116 (21.9) |  |
| Number of passes, median (IQR) | 2.0 (1.0-2.0) | 3.0 (2.0-4.0) | 2.0 (1.0-2.0) | <0.001 |
| Onset-to-recanalization, min, median (IQR) | 500.0 (347.5-756.0) | 455.0 (328.5-706.0) | 505.0 (350.0-764.3) | 0.149 |
| Rescue therapy |  |  |  | 0.076 |
| No | 464 (77.6) | 59 (86.8) | 405 (76.4) |  |
| Yes | 134 (22.4) | 9 (13.2) | 125 (23.6) |  |
| **mTICI Grade, n (%)** |  |  |  | 0.025 |
| 0 | 26 (4.3) | 1 (1.5) | 25 (4.7) |  |
| 1 | 20 (3.3) | 4 (5.9) | 16 (3.0) |  |
| 2a | 41 (6.9) | 7 (10.3) | 34 (6.4) |  |
| 2b | 208 (34.8) | 32 (47.1) | 176 (33.2) |  |
| 3 | 303 (50.7) | 24 (35.3) | 279 (52.6) |  |
| **Laboratory Parameters** |  |  |  |  |
| NLR, median (IQR) | 3.97 (3.38-4.85) | 4.31 (3.77-5.52) | 3.91 (3.32-4.81) | 0.005 |
| PLR, median (IQR) | 106.36 (58.24-162.98) | 124.68 (68.67-185.88) | 104.55 (57.74-160.89) | 0.089 |
| SIRI, 10<sup>9</sup>, median (IQR) | 2.59 (1.50-5.68) | 2.57 (1.61-6.25) | 2.59 (1.50-5.54) | 0.555 |
| D-dimer, median (IQR) | 0.32 (0.24-0.68) | 0.37 (0.26-1.42) | 0.32 (0.24-0.53) | 0.036 |
| CRP, median (IQR) | 5.06 (1.24-13.08) | 3.97 (0.98-14.57) | 5.11 (1.24-13.01) | 0.813 |
| Admission glucose, mmol/L, median (IQR) | 7.50 (6.00-10.30) | 7.75 (6.15-12.17) | 7.45 (6.00-10.30) | 0.368 |

Abbreviations as in Table 1.

For external validation, we identified 138 patients from three independent sources (Figure S1B). HT occurred in 54 patients (39.1%), and their baseline characteristics are summarized in Table S2.

### Independent Predictors of HT

Univariable analysis identified nine candidate variables that were entered into the multivariable logistic regression model (P<0.10; Table 1). In multivariable analysis, five variables were retained as independent predictors of HT (Table 3): admission NIHSS (OR, 1.055 per point; 95% CI, 1.019-1.092; P=0.002), atrial fibrillation (OR, 2.279; 95% CI, 1.325-3.908; P=0.003), alcohol consumption (OR, 1.725; 95% CI, 1.162-2.563; P=0.007), number of thrombectomy passes (OR, 1.965 per pass; 95% CI, 1.608-2.422; P<0.001), and mTICI grade (overall P=0.003). The mTICI grade showed a non-linear, inverted U-shaped association with HT risk. Compared with the worst reperfusion (Grade 0), the odds of HT peaked at partial recanalization (Grade 2a: OR, 7.410 [95% CI, 2.071-31.975]; P=0.004; Grade 2b: OR, 7.502 [95% CI, 2.430-29.248]; P=0.001) and declined at complete recanalization (Grade 3: OR, 4.972 [95% CI, 1.594-19.542]; P=0.011).

**Table 3.** Logistic Regression Analysis for Predictors of Hemorrhagic Transformation in the Derivation Cohort.

| Variable | Univariate Regression Model |  | Multivariable Regression Model |  |  |
| --- | --- | --- | --- | --- | --- |
| | OR (95% CI) | P Value | $\beta$ | OR (95% CI) | P Value |
| Age | 0.982 (0.947-1.019) | 0.326 | — | — | — |
| Admission NIHSS | 1.048 (1.011-1.087) | 0.011 | 0.053 | 1.055 (1.019-1.092) | 0.002 |
| ASPECTS | 0.925 (0.826-1.036) | 0.174 | — | — | — |
| Number of passes | 1.897 (1.543-2.350) | <0.001 | 0.676 | 1.965 (1.608-2.422) | <0.001 |
| Atrial fibrillation | 2.369 (1.361-4.115) | 0.002 | 0.824 | 2.279 (1.325-3.908) | 0.003 |
| Alcohol consumption | 1.770 (1.186-2.646) | 0.005 | 0.545 | 1.725 (1.162-2.563) | 0.007 |
| Rescue therapy | 1.003 (0.607-1.631) | 0.992 | — | — | — |
| Collateral circulation |  |  | — | — | — |
| — Linear trend | 0.844 (0.612-1.166) | 0.301 |  |  |  |
| — Quadratic trend | 1.486 (1.026-2.184) | 0.040 |  |  |  |
| mTICI Grade |  | 0.001 |  |  | 0.003 |
| 1 |  |  | 1.680 | 5.344 (1.224-26.653) | 0.031 |
| 2a |  |  | 2.000 | 7.410 (2.071-31.975) | 0.004 |
| 2b |  |  | 2.020 | 7.502 (2.430-29.248) | 0.001 |
| 3 |  |  | 1.600 | 4.972 (1.594-19.542) | 0.011 |
| — Linear trend | 3.388 (1.500-8.681) | 0.006 |  |  |  |
| — Quadratic trend | 0.275(0.118-0.589) | 0.001 |  |  |  |
| — Cubic trend | 1.472 (0.685-3.165) | 0.317 |  |  |  |
| — Quartic trend | 0.853 (0.420-1.737) | 0.659 |  |  |  |

### The NO-PAIN Score and Risk Stratification

Based on the five independent predictors, the final prediction model was established and used to derive the NO-PAIN Score (Table 4). The total score ranged from 9.8 to 53.4. The Spearman correlation between the total score and the predicted probability from the regression model was 0.9998. Patients were stratified into three risk groups according to the total score: low risk (≤26.2 points), moderate risk (26.2-32.2 points), and high risk (>32.2 points). The rates of HT across these groups were 13.9%, 25.1%, and 50.3%, respectively (Table S3).

**Table 4.** Simplified Point-Score System for Predicting Hemorrhagic Transformation.

| Variable | Points |
| --- | --- |
| Admission NIHSS (per point) | 0.4 |
| Number of passes (per pass) | 5 |
| Alcohol consumption | 4 |
| Atrial fibrillation | 6 |
| mTICI Grade |  |
| 0 | 0 |
| 1 | 12 |
| 2a | 15 |
| 2b | 15 |
| 3 | 12 |

The point-score system was derived from the $\beta$ coefficients of the final parsimonious model, with the number of thrombectomy passes serving as the reference variable (assigned 5 points per pass). The total score ranges from 9.8 to 53.4. The Spearman correlation between the total score and the predicted probability was 0.9998.

### Internal and External Validation

The final model demonstrated acceptable discrimination, with a C-index of 0.737 (95% CI, 0.693-0.782) in the derivation cohort (Figure 1A). After 1000 bootstrap resamples, the optimism-corrected C-index was 0.748 (95% CI, 0.705-0.785). The Hosmer-Lemeshow test showed good calibration (P=0.520), and the calibration plot confirmed close agreement between prediction and observation, with a mean absolute error of 0.014 (Figure 2A). Decision curve analysis demonstrated that the model yielded a higher net benefit than the treat-all or treat-none strategies across a threshold probability range of 3% to 50% (Figure 3A). The net benefit of the treat-all strategy fell below zero at the observed HT prevalence of 29.4%.

**Figure 1.**
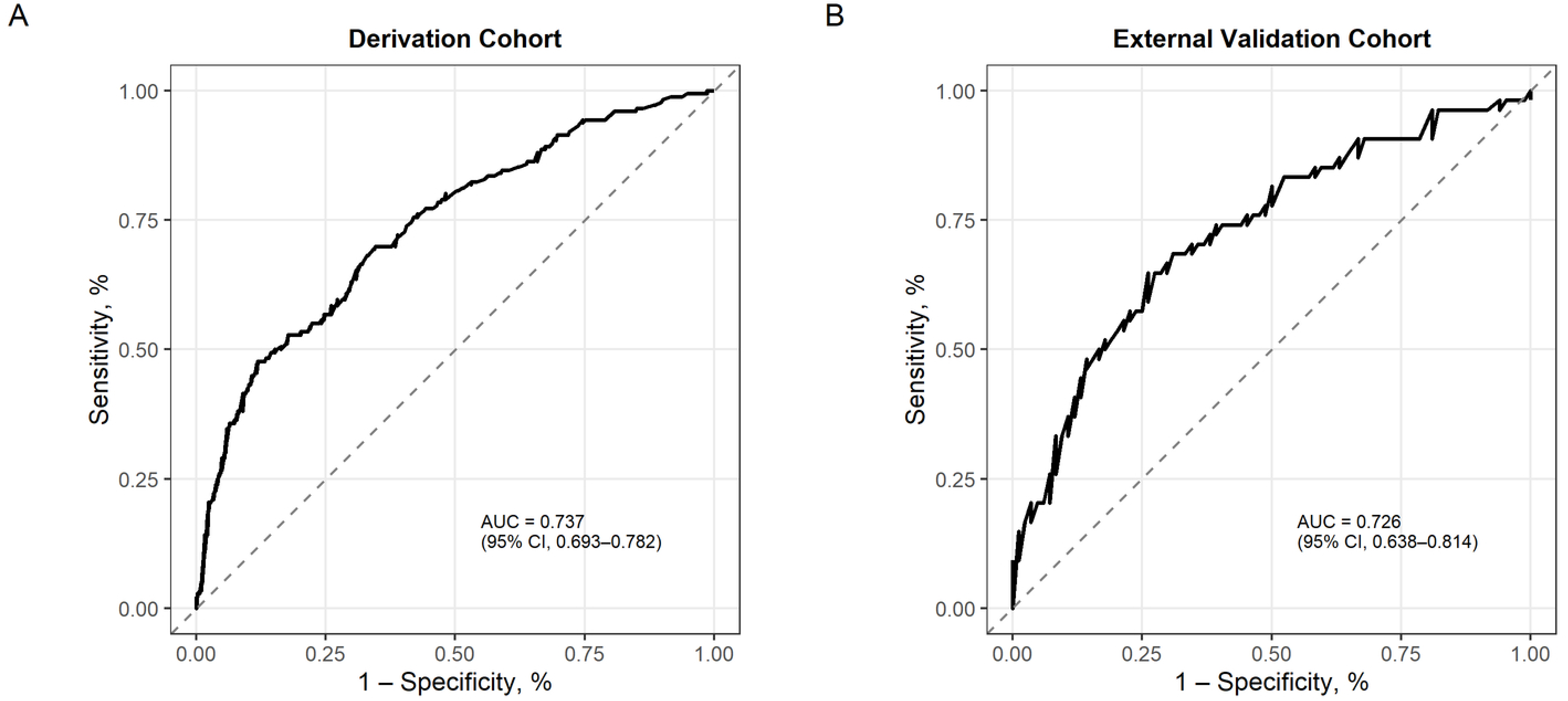
ROC curves for the HT prediction model. (A) Derivation cohort: AUC = 0.737 (95% CI, 0.693 - 0.782); optimism-corrected AUC after 1000 bootstrap resamples = 0.748 (95% CI, 0.705-0.785). (B) External validation cohort: AUC = 0.726 (95% CI, 0.638-0.814). AUC indicates area under the curve. The dashed diagonal line represents the reference line of no discrimination.

**Figure 2.**
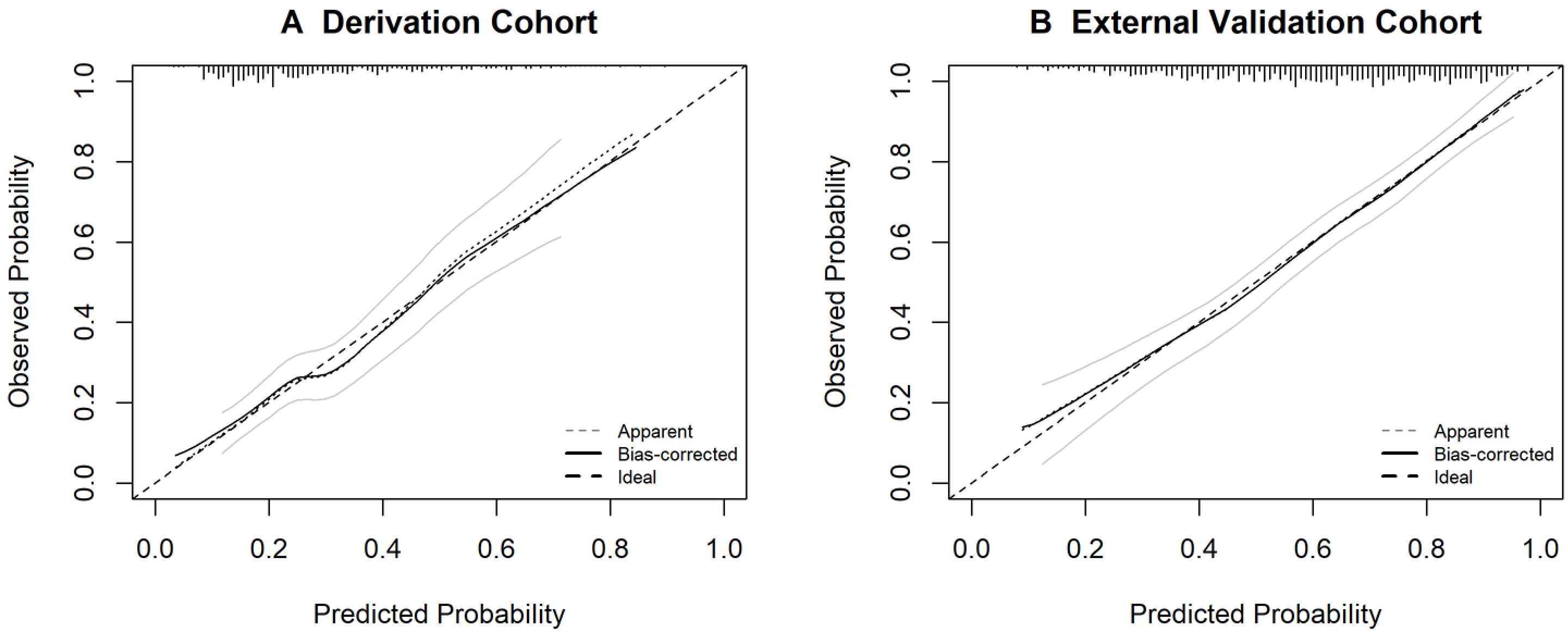
Calibration plots for the prediction model. (A) Derivation cohort: Hosmer-Lemeshow P = 0.520, mean absolute error = 0.014. (B) External validation cohort: Hosmer-Lemeshow P = 0.789, mean absolute error = 0.007. The solid black line represents the smoothed calibration curve. The gray band indicates the pointwise 95% confidence interval derived from 200 bootstrap resamples. The dashed diagonal line represents perfect calibration.

**Figure 3.**
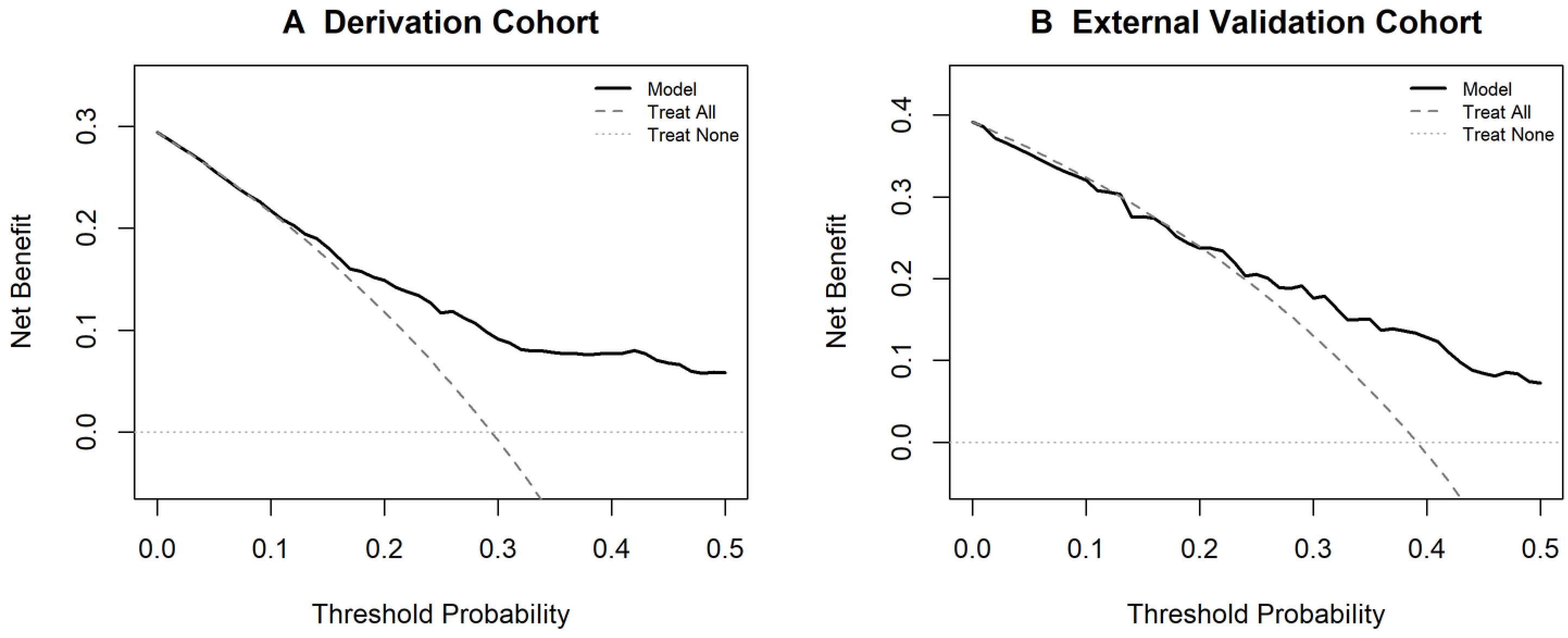
Decision curve analysis for the prediction model. (A) Derivation cohort: the model yielded a higher net benefit than the treat-all or treat-none strategies across a threshold probability range of 3% to 50%. (B) External validation cohort: the model outperformed treat-all across a threshold range of 13% to 50%. The net benefit of the treat-all strategy fell below zero at a prevalence of 29.4% in the derivation cohort and 39.1% in the external cohort.

In the external validation cohort, the model maintained acceptable discrimination, with a C-index of 0.726 (95% CI, 0.638-0.814; Figure 1B). Calibration was also acceptable (Hosmer-Lemeshow P=0.789; mean absolute error, 0.007; Figure 2B). Decision curve analysis showed that the model outperformed the treat-all strategy across a threshold range of 13% to 50%, and the net benefit of treat-all fell below zero at the observed HT prevalence of 39.1% (Figure 3B).

### Subgroup Analysis of sICH

In the exploratory subgroup analysis for sICH (68/598, 11.4%), admission NIHSS (OR, 1.067 per point; 95% CI, 1.018-1.118; P=0.007) and the number of thrombectomy passes (OR, 2.165 per pass; 95% CI, 1.681-2.814; P<0.001) remained significant predictors. Better collateral circulation emerged as an independent protective factor for sICH (OR, 0.490 per grade; 95% CI, 0.259-0.915; P=0.026), which was not observed in the overall HT model. A comparison of independent predictors between HT and sICH is presented in Table 5.

**Table 5.** Independent Predictors of HT vs. sICH.

| Variable | HT (n=176) |  | sICH (n=68) |  |
| --- | --- | --- | --- | --- |
|  | OR (95% CI) | P Value | OR (95% CI) | P Value |
| **Common Predictors** |  |  |  |  |
| Admission NIHSS | 1.055 (1.019-1.092) | 0.002 | 1.067 (1.018-1.118) | 0.007 |
| Number of passes | 1.965 (1.608-2.422) | <0.001 | 2.165 (1.681-2.814) | <0.001 |
| **HT-Specific Predictors** |  |  |  |  |
| mTICI grade (overall) | — | 0.003 | — | NS |
| Alcohol consumption | 1.725 (1.162-2.563) | 0.007 | — | NS |
| Atrial fibrillation | 2.279 (1.325-3.908) | 0.003 | — | NS |
| **sICH-Specific Predictor** |  |  |  |  |
| Collateral circulation | — | NS | — | 0.003 |

HT indicates hemorrhagic transformation; sICH, symptomatic intracranial hemorrhage; OR, odds ratio; CI, confidence interval; NIHSS, National Institutes of Health Stroke Scale; mTICI, modified Thrombolysis in Cerebral Infarction; NS, not significant ( $P \geq 0.05$ ).

## Discussion

In this multicenter retrospective study, we developed and externally validated a prediction model for HT after EVT in young adults with acute anterior circulation large vessel occlusion. Five independent predictors were identified—admission NIHSS, number of thrombectomy passes, atrial fibrillation, alcohol consumption, and mTICI grade—and built into a simplified score, termed the NO-PAIN Score. The score demonstrated satisfactory predictive performance in both the derivation and external validation cohorts, with acceptable calibration across samples. These findings suggest that the NO-PAIN Score may serve as a practical tool for individualized risk stratification in this patient population.

Previous prediction models for HT after EVT have focused predominantly on sICH, owing to its strong prognostic impact.^12–14^ However, accumulating evidence suggests that asymptomatic HT also holds prognostic significance. A pooled analysis of the RESCUE BT and DEVT trials found that asymptomatic intracranial hemorrhage was independently associated with less neurologic improvement and greater 90-day disability.^19^ Moreover, asymptomatic HT has been shown to be independently associated with a greater need for inpatient rehabilitation and a reduced likelihood of functional independence in recent studies.^9,11^ In young adults undergoing EVT, prediction models for HT—not simply sICH—have rarely been developed. Therefore, we developed our prediction model with HT as the primary endpoint.

Among the five predictors of the final model, the number of thrombectomy passes was the strongest. Multiple retrieval attempts have been consistently associated with increased endothelial disruption, blood-brain barrier injury, and a higher risk of both sICH and any HT.^12,20–22^ Atrial fibrillation was the second strongest predictor, which may reflect the greater thrombus burden and the technically more demanding retrieval associated with cardioembolic stroke.^20,21^ A history of alcohol consumption was also independently associated with HT. Chronic alcohol use has been linked to endothelial dysfunction and impaired coagulation, which may predispose to hemorrhagic transformation, and prior thrombectomy studies have reported similar associations.^1,22^ Admission NIHSS, a classic marker of initial stroke severity, showed a modest effect, consistent with its established role across numerous prediction scores.^7,12,20, 24^ Finally, the mTICI grade displayed a distinctly non-linear, inverted U-shaped relationship with HT risk, an observation that departs from the traditional assumption of a monotonic association between recanalization success and hemorrhage.

A notable observation in this study is the non-linear, inverted U-shaped relationship between mTICI grade and HT risk. The highest risk was observed at partial recanalization(Grade 2a/2b), rather than at either the worst or best reperfusion. This pattern is consistent with the concept of reperfusion injury, wherein incomplete flow restoration causes greater blood-brain barrier disruption than either persistent occlusion or full recanalization.^25,26^ Although a similar non-linear pattern has been noted in earlier studies, it has rarely been modeled explicitly, and several widely used scores treat recanalization as either a binary variable or a linear term.^27–29^ Clinically, the inverted U-shaped pattern underscores the importance of achieving complete(Grade 3) rather than partial recanalization. Patients with incomplete recanalization may therefore warrant closer post-procedural monitoring.

The discriminative performance of the NO-PAIN Score was similar to the c-statistics reported for the ASIAN score (0.77) and the TREAT-AIS score (approximately 0.80), although these instruments were designed to predict sICH rather than any radiographic HT and were not specifically developed for young adults.^12,14^ The utility of the NO-PAIN Score lies primarily in its simplicity—all constituent variables are routinely available in clinical practice, and the predominantly integer-based scoring allows rapid bedside calculation.

The risk factor profiles for HT and sICH shared several predictors but were not entirely concordant. Admission NIHSS and the number of thrombectomy passes emerged as predictors of both outcomes, in line with their established roles as markers of initial stroke severity and procedural complexity.^7,20^ However, alcohol consumption, and atrial fibrillation were associated only with HT, suggesting that these factors may contribute primarily to milder, asymptomatic hemorrhages rather than to sICH. Conversely, better collateral circulation was independently protective against sICH but did not enter the HT model. This observation is consistent with previous studies showing that robust collaterals reduce the risk of symptomatic hemorrhage by limiting infarct progression and attenuating reperfusion injury.^23^ This distinction may reflect a two-stage process, in which the occurrence of HT and its progression to symptomatic deterioration are influenced by different sets of factors. This hypothesis, however, requires further investigation. These insights may help guide post-procedural monitoring and decisions on early antithrombotic therapy.

## Limitations

Several limitations of this study should be noted. The retrospective design carries inherent risks of selection bias. The external validation cohort was derived from three independent data sources with different study designs and inclusion criteria, which may have introduced heterogeneity. In addition, the analysis was confined to anterior circulation occlusion. Posterior circulation stroke differs from anterior circulation stroke in etiology, technical aspects of thrombectomy, and HT mechanisms.^30,31^ By restricting the cohort to anterior circulation occlusion, we sought to enhance the homogeneity of the study population.

## Conclusions

In this multicenter study, a prediction model for HT after EVT was developed and externally validated in young adults with acute anterior circulation large vessel occlusion. Five independent predictors were incorporated into a simplified bedside score, the NO-PAIN Score, which may serve as a practical tool for individualized risk stratification in this population.

## Author Contributions

Dr X.L. and W.Z. contributed to the study conception and design, and were responsible for critical review of the manuscript and its final editing. Q.L. drafted the article and participated in data analysis and interpretation. Q.L., K.Y. and A.L. contributed to the study design and participated in methodological operations. Z. W., Y.L., G. X., W. L., Z. Z., D.Y., K.H., C.C., W. D. and L. P. participated in data collection. All authors have read and approved the final version of the article.

## Supplemental Material

Tables S1-S3

Figure S1

## Non-standard Abbreviations and Acronyms

HT: hemorrhagic transformation
EVT: endovascular thrombectomy
sICH: symptomatic intracranial hemorrhage
NIHSS: National Institutes of Health Stroke Scale
mRS: modified Rankin Scale
ASPECTS: Alberta Stroke Program Early CT Score
mTICI: modified Thrombolysis in Cerebral Infarction
NLR: neutrophil-to-lymphocyte ratio
PLR: platelet-to-lymphocyte ratio
SIRI: systemic inflammatory response index
ASITN/SIR: American Society of Interventional and Therapeutic Neuroradiology/Society of Interventional Radiology
TOAST: Trial of ORG 10172 in Acute Stroke Treatment
TIA: transient ischemic attack

## Data Availability

The data that support the findings of this study are available from the corresponding author upon reasonable request. The data are not publicly available because they contain information that could compromise the privacy of research participants and the sharing of data requires additional approval from the institutional review boards of the participating centers.

## References

1. Ekker MS, Boot EM, Singhal AB, et al. Epidemiology, aetiology, and management of ischaemic stroke in young adults. Lancet Neurol. 2018;17(9):790–801. doi: 10.1016/S1474-4422(18)30233-3.

2. Boot E, Ekker MS, Putaala J, et al. Ischaemic stroke in young adults: a global perspective. J Neurol Neurosurg Psychiatry. 2020;91(4):411–417. doi: 10.1136/jnnp-2019-322424.

3. Synhaeve NE, Arntz RM, Maaijwee NA, et al. Poor long-term functional outcome after stroke among adults aged 18 to 50 years: Follow-Up of Transient Ischemic Attack and Stroke Patients and Unelucidated Risk Factor Evaluation (FUTURE) study. Stroke. 2014;45(4):1157–1160. doi: 10.1161/STROKEAHA.113.004411.

4. Brouwer J, Smeijers AS, Mulder MJ, et al. Endovascular thrombectomy in young patients with stroke: a MR CLEAN registry study. Stroke. 2022;53(1):34–42. doi: 10.1161/STROKEAHA.120.034033.

5. Nicolini E, Saia V, Lorenzano S, et al. Mechanical thrombectomy in young patients with large vessel occlusion-related ischemic stroke: Data from the Italian Registry of Endovascular Treatment in Acute Stroke. Eur J Neurol. 2023;30(12):3751–3760. doi: 10.1111/ene.16035.

6. Lee YB, Yoon W, Lee YY, et al. Predictors and impact of hemorrhagic transformations after endovascular thrombectomy in patients with acute large vessel occlusions. J Neurointerv Surg. 2019;11(5):469–473. doi: 10.1136/neurintsurg-2018-014080.

7. Kaesmacher J, Kaesmacher M, Maegerlein C, et al. Hemorrhagic transformations after thrombectomy: risk factors and clinical relevance. Cerebrovasc Dis. 2017;43:294–304. doi: 10.1159/000460265.

8. Li W, Xing X, Wen C, et al. Risk factors and functional outcome were associated with hemorrhagic transformation after mechanical thrombectomy for acute large vessel occlusion stroke. J Neurosurg Sci. 2023;67(5):585–590. doi: 10.23736/S0390-5616.20.05141-3.

9. Guasch-Jiménez M, Ezcurra Díaz G, Lambea-Gil Á, et al. Influence of Asymptomatic Hemorrhagic Transformation After Endovascular Treatment on Stroke Outcome: A Population-Based Study. Neurology. 2025;104(9):e213509. doi: 10.1212/WNL.0000000000213509.

10. Suzuki K, Katano T, Numao S, et al. The effect of asymptomatic intracranial hemorrhage after mechanical thrombectomy on clinical outcome. J Neurol Sci. 2024;457:122868. doi: 10.1016/j.jns.2024.122868.

11. Hung A, et al. Clinically Asymptomatic Hemorrhagic Conversion Is Associated with Need for Inpatient Rehabilitation After Mechanical Thrombectomy for Anterior Circulation Ischemic Stroke. World Neurosurg. 2024;186:e181–e190. doi: 10.1016/j.wneu.2024.03.102.

12. Zhang X, Xie Y, Wang H, et al. Symptomatic Intracranial Hemorrhage After Mechanical Thrombectomy in Chinese Ischemic Stroke Patients: The ASIAN Score. Stroke. 2020;51(9):2690–2696. doi: 10.1161/STROKEAHA.120.030173.

13. Cappellari M, Mangiafico S, Saia V, et al. IER-SICH Nomogram to Predict Symptomatic Intracerebral Hemorrhage After Thrombectomy for Stroke. Stroke. 2019;50(4):909–916. doi: 10.1161/STROKEAHA.

14. Chen JH, Su IC, Lu YH, et al. Predictive Modeling of Symptomatic Intracranial Hemorrhage Following Endovascular Thrombectomy: Insights From the Nationwide TREAT-AIS Registry. J Stroke. 2025;27(1):85–94. doi: 10.5853/jos.2024.04119.

15. von Elm E, Altman DG, Egger M, et al. The Strengthening the Reporting of Observational Studies in Epidemiology (STROBE) statement: guidelines for reporting observational studies. Lancet. 2007;370(9596):1453–1457. doi: 10.1016/S0140-6736(07)61602-X.

16. Zi W, Wang H, Yang D, et al. Clinical effectiveness and safety outcomes of endovascular treatment for acute anterior circulation ischemic stroke in China. Cerebrovasc Dis. 2017;44(5-6):248–258. doi: 10.1159/000478667.

17. Von Kummer R, Broderick JP, Campbell BC, et al. The Heidelberg bleeding classification: classification of bleeding events after ischemic stroke and reperfusion therapy. Stroke. 2015;46(10):2981–2986. doi: 10.1161/STROKEAHA.115.010049.

18. Zaidat OO, Yoo AJ, Khatri P, et al. Recommendations on angiographic revascularization grading standards for acute ischemic stroke: a consensus statement. Stroke. 2013;44(9):2650–63. doi: 10.1161/STROKEAHA.113.001972.

19. Cai L, Jin W, Albers GW, et al. Association Between Asymptomatic Intracranial Hemorrhage and Outcomes After Thrombectomy: A Pooled Analysis of the RESCUE BT and DEVT Trials. Neurol Clin Pract. 2025;15(4):e200500. doi: 10.1212/CPJ.0000000000200500.

20. Hao Y, Yang D, Wang H, et al. Predictors for symptomatic intracranial hemorrhage after endovascular treatment of acute ischemic stroke. Stroke. 2017;48(5):1203–1209. doi: 10.1161/STROKEAHA.116.016368.

21. Sun J, Lam C, Christie L, et al. Risk factors of hemorrhagic transformation in acute ischaemic stroke: A systematic review and meta-analysis. Front Neurol. 2023;14:1079205. doi: 10.3389/fneur.2023.1079205.

22. Shi Z, Luo G, Huo X, et al. Predictors of parenchymal hemorrhage after endovascular treatment in large core ischemic stroke: a post-hoc analysis of the ANGEL-ASPECT trial. J Neurointerv Surg. 2026 Mar 13;18(4):942–949. doi: 10.1136/jnis-2025-023285.

23. Qian J, Lu F, Zhang W, et al. A meta-analysis of collateral status and outcomes of mechanical thrombectomy. Acta Neurol Scand. 2020 Sep;142(3):191–199. doi: 10.1111/ane.13255.

24. Lou M, Safdar A, Mehdiratta M, et al. The HAT Score: a simple grading scale for predicting hemorrhage after thrombolysis. Neurology. 2008;71(18):1417–1423. doi: 10.1212/01.wnl.0000330297.58334.dd.

25. Khatri R, McKinney AM, Swenson B, et al. Blood-brain barrier, reperfusion injury, and hemorrhagic transformation in acute ischemic stroke. Neurology. 2012;79(13 Suppl 1):S52–7. doi: 10.1212/WNL.0b013e3182697e70.

26. Warach S, Latour LL. Evidence of reperfusion injury, exacerbated by thrombolytic therapy, in human focal brain ischemia using a novel imaging marker of early blood-brain barrier disruption. Stroke. 2004;35(11 Suppl 1):2659–61. doi: 10.1161/01.STR.0000144051.32131.09.

27. Bang OY, Saver JL, Kim SJ, et al. Collateral flow averts hemorrhagic transformation after endovascular therapy for acute ischemic stroke. Stroke. 2011;42(8):2235–2239. doi: 10.1161/STROKEAHA.110.604603.

28. Flint AC, Cullen SP, Faigeles BS, et al. THRIVE score predicts outcomes with a third-generation endovascular stroke treatment device. Stroke. 2013;44(12):3370–5. doi: 10.1161/STROKEAHA.113.002796.

29. Tanaka K, Brown S, Goyal M, et al. HERMES-24 score derivation and validation for simple and robust outcome prediction after large vessel occlusion treatment. Stroke. 2024;55(8):1982–1990. doi: 10.1161/STROKEAHA.123.045871.

30. Mbroh J, Poli K, Tünnerhoff J, Gomez-Exposito A, et al. Comparison of risk factors, safety, and efficacy outcomes of mechanical thrombectomy in posterior vs. anterior circulation large vessel occlusion. Front Neurol. 2021;12:687134. doi: 10.3389/fneur.2021.687134.

31. Hendrix P, Killer-Oberpfalzer M, Broussalis E, et al. Mechanical thrombectomy for anterior versus posterior circulation large vessel occlusion stroke. World Neurosurg. 2022;158:e416–e422. doi: 10.1016/j.wneu.2021.10.187.

